# Genetic and epigenetic signatures associated with plasma oxytocin levels in children and adolescents with autism spectrum disorder

**DOI:** 10.1101/2022.05.26.22275542

**Authors:** Stephen K Siecinski, Stephanie N Giamberardino, Marina Spanos, Annalise C Hauser, Jason R Gibson, Tara Chandrasekhar, M D Pilar Trelles, Carol M Rockhill, Michelle L Palumbo, Allyson Witters Cundiff, Alicia Montgomery, Paige Siper, Mendy Minjarez, Lisa A Nowinski, Sarah Marler, Lydia C Kwee, Lauren C Shuffrey, Cheryl Alderman, Jordana Weissman, Brooke Zappone, Jennifer E Mullett, Hope Crosson, Natalie Hong, Sheng Luo, Lilin She, Manjushri Bhapkar, Russell Dean, Abby Scheer, Jacqueline L Johnson, Bryan H King, Christopher J McDougle, Kevin B Sanders, Soo-Jeong Kim, Alexander Kolevzon, Jeremy Veenstra-VanderWeele, Elizabeth R Hauser, Linmarie Sikich, Simon G Gregory

## Abstract

Oxytocin (OT), the brain’s most abundant neuropeptide, plays an important role in social salience and motivation. Clinical trials of the efficacy of OT in autism spectrum disorder (ASD) have reported mixed results due in part to ASD’s complex etiology. We hypothesized that genetic and epigenetic variation contribute to variable endogenous OT levels that modulate sensitivity to OT therapy. To test this hypothesis, we integrated genome-wide profiles of DNA-methylation, transcriptional activity, and genetic variation with plasma OT levels in 290 participants with ASD enrolled in a randomized controlled trial of OT. Our analysis shows subtle, but statistically significant association of plasma OT levels with peripheral transcriptional activity and DNA-methylation profiles across several annotated gene sets. We also identified genetic variants with novel association with plasma OT, several of which reside in known ASD risk genes. These findings broaden our understanding of the effects of the peripheral oxytocin system and provide novel genetic candidates for future studies to decode the complex etiology of ASD and its interaction with OT signaling and OT-based interventions.

## INTRODUCTION

Oxytocin (OT) is the most abundant neuropeptide in the human brain and functions as a neurotransmitter, neuromodulator, and hormone with actions throughout the body ^1-3^. In the brain, oxytocin is thought to contribute to social attachment, affiliation, and sexual behavior at least partially through the modulation of mesolimbic dopaminergic neural circuits ^3^. This is further supported by a recent brain-wide voxel-based transcriptional and fMRI meta-analysis that showed region-specific overlap of oxytocin activity with dopaminergic and cholinergic genes in brain regions associated with appetitive and aversive emotional states ^4^. Oxytocin exerts these neuromodulatory effects via somatodendritic release from magnocellular neurons of the supraoptic and paraventricular nuclei (SON and PVN, respectively) of the hypothalamus and subsequent binding to its widely distributed receptor, OXTR ^5-13^. Collectively, the broad neuromodulatory and prosocial effects of oxytocin make it an attractive candidate for therapeutic intervention targeting social difficulties, particularly in ASD ^14^.

Clinical trials of oxytocin in humans aim to improve social deficits in a broad range of conditions including addiction, eating disorders, schizophrenia, and ASD. To date, 39 clinical trials have been conducted to evaluate the safety and efficacy of intranasal oxytocin in the context of ASD alone, with over 240 trials across all diagnostic categories. Oxytocin trials in ASD have had mixed results, with some showing no improvement in core social deficits ^15-17^, including our own study ^18^, while others have shown improvements in specific populations ^19^ or in individuals with co-occurring intellectual disability ^20^. Important to assessing oxytocin treatment efficacy and mode of action in the context of behavioral phenotypes is an individual’s baseline levels of endogenously produced oxytocin. Recent studies have shown that baseline oxytocin levels may play an important role in the presentation of ASD ^21, 22^ and social behaviors more broadly ^23-29^ and may be influenced by putative genetic factors ^30^.

Genetic and epigenetic studies have previously evaluated components of the oxytocin system in relation to social behavior and autism. Candidate gene studies have reported associations between polymorphisms in oxytocin system genes and social phenotypes, including in ASD; although this has not been reported in genome-wide association studies (GWAS) ^31-42^. We previously reported that differential methylation of the CpG-island (CGI) of the *OXTR* promoter is associated with ASD ^43^, while additional studies have identified this association with various other psychosocial phenotypes ^44-48^. This suggests that peripheral measurements of *OXTR* methylation could serve as a biomarker to identify OT pathway dysregulation and may correlate with baseline levels of the ligand, which has a short peripheral half-life. Few studies, and none in ASD, have previously performed genome-wide analyses in relation to plasma oxytocin levels^49^.

This study investigates the genetic, epigenetic, and transcriptomic variation associated with plasma levels of oxytocin in biospecimens from 175 participants from the Study of Oxytocin in Autism and Reciprocal Social Behaviors (SOARS-B) trial ^18^, and provide the most comprehensive genetic and epigenetic analysis of the peripheral oxytocin system to date.

## METHODS and MATERIALS

### Study Population and Sample Selection

A full description of the SOARS-B clinical trial and study population has been previously reported ^18, 50^. Briefly, 355 children and adolescents were screened for study inclusion, with 290 individuals with ASD enrolled and randomized 1:1 to intranasal placebo or oxytocin spray in a multi-site clinical trial. Participants who passed screening and had available DNA or RNA samples were eligible for inclusion in the current study (Table 1). Participant samples were considered for inclusion for each modality independently (Figure 1) with blood drawn at baseline/screening and weeks 8, 24, 36, and 48. To maximize the sample size for the genome-wide association study (GWAS), DNA samples were genotyped from any available timepoint. Samples for other genomic modalities were restricted to those obtained at screening and/or baseline study visits, collectively referred to as “pre-treatment” samples. Notably, because gene expression profiling was designed to profile participants longitudinally, only participants with a viable RNA sample available from at least 2 different study visits were included in this dataset (defined as containing at least 0.05 µg). SOARS-B sample collection was reviewed and approved by the institutional review board at each site. Written parental consent and participant assent (when clinically appropriate) was obtained for all participants and the UNC Tracs Data Safety Monitoring Board (DSMB) reviewed the safety data throughout the study. Analyses of the samples were carried out under two Duke IRBs (Pro00063950, Pro00016404).

**Table 1:** Population characteristics for participants with pre-treatment plasma oxytocin and at least one other dataset (after quality control).

| Characteristic | Analysis Population (n=175) |
| --- | --- |
| Mean age in years at baseline (SD) | 10.74 (3.91) |
| Female sex (%) | 25 ( 14.3) |
| Self-Report Racial Group (%) |  |
| White | 125 (71.4) |
| Black/African America | 15 (8.6) |
| Asian | 18 (10.3) |
| More than one race | 15 (8.6) |
| Not reported | 2 (1.1) |
| Hispanic/Latino (%) | 16 (9.1) |
| Low Functioning Group (%) | 91 (52.0) |
| Study Site (%) |  |
| Duke (previously UNC) | 50 (28.6) |
| Mt. Sinai School of Medicine | 20 (11.4) |
| Massachusetts General Hospital | 15 (8.6) |
| Vanderbilt | 51 (29.1) |
| Seattle Children's Research Institute | 39 (22.3) |
| Post-QC Dataset Availability (%) |  |
| Illumina Omni 2.5 genotypes | 175 (100.0) |
| Targeted genotypes | 175 (100.0) |
| CNVs | 157 (89.7) |
| Illumina HT-12 gene expression | 149 (85.1) |
| Illumina EPIC methylation | 174 (99.4) |
| OXTR targeted methylation | 172 (98.3) |

**Figure 1.**
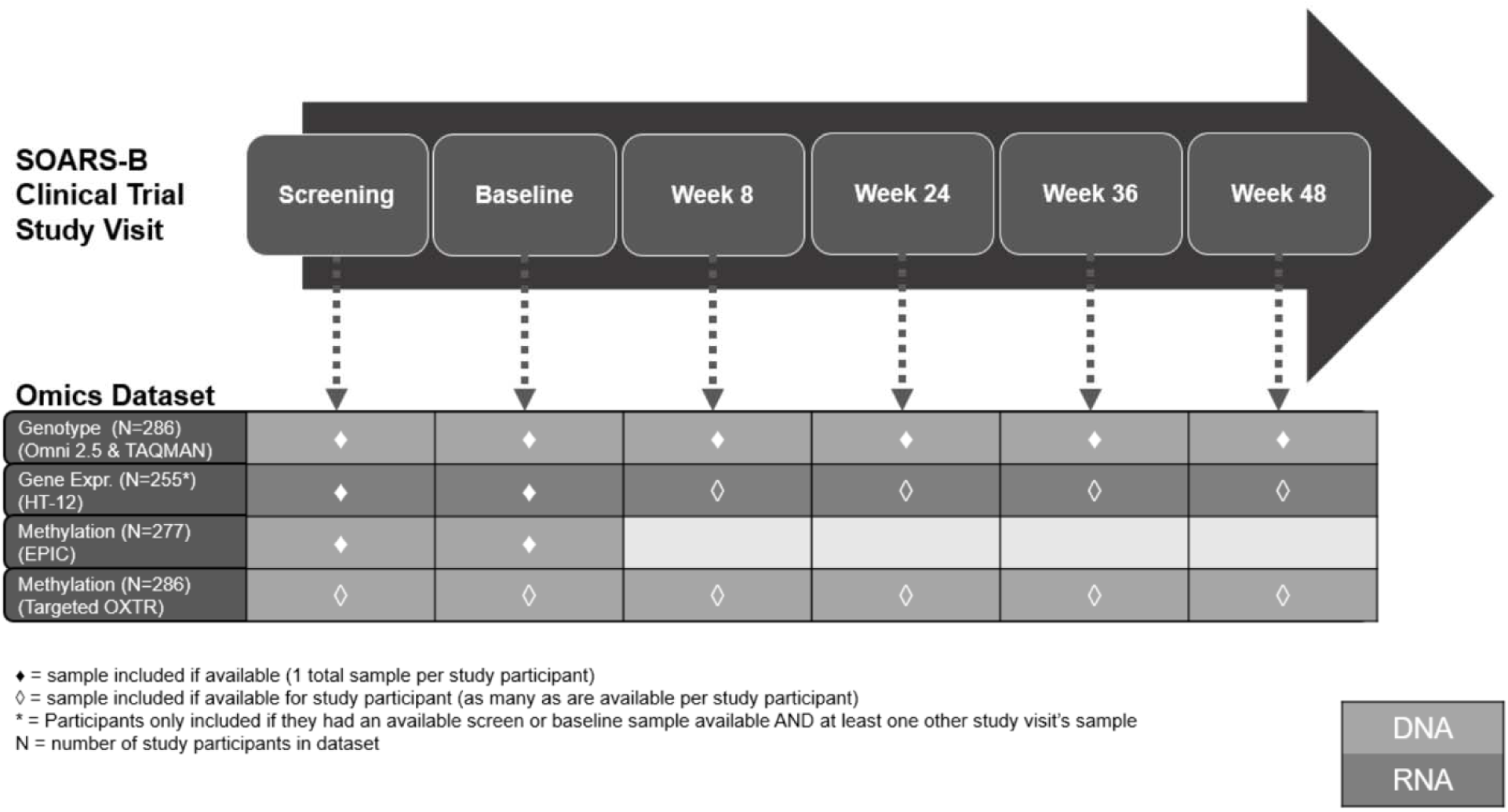
Participant and sample inclusion for each modality tested within the study

### Plasma Oxytocin

Plasma was extracted from 7 mL of blood, and plasma OT levels were quantified in 96-well batches using the Enzo Oxytocin ELISA kit (CAT: ADI-901-153A) with the standard protocol at the University of North Carolina, as previously described ^51^. Each sample was quantified (pg/mL) on a standard curve, and mean plasma OT values were included if more than one pre-treatment measurement was available for a given sample. Sample measurements that could not be validated after quality control were excluded from downstream analyses (N=8 across all samples with measurements), as well as samples from the Columbia study site due to systematic differences in the OT measures (Supplementary Figure 1). For all subsequent analyses, natural log-transformed pre-treatment plasma OT values were used.

### Genome-Wide Genotyping, Expression and Methylation Profiling

The hg19 (GRCh37) reference genome was used for all quality control and analytical operations described in the genotype, gene expression, and methylation profiling methods.

Genome-wide genotype profiling was performed using the Illumina Infinium Omni2.5 BeadChip array for 286 study participants and 6 Centre d’Etude du Polymorphism Humain (CEPH) samples from Coriell, including one parent-offspring quad. Data processing and quality control were performed within Illumina GenomeStudio and PLINK (v1.9) ^52^. Briefly, variants were excluded if they were classified as insertions or deletions and for poor genotyping calling, coordinate mapping issues, discordance between identical samples, Mendelian errors, haploid heterozygosity, and excessive missingness or monomorphism within our dataset. Samples were excluded for genotypic-phenotypic sex-discrepancies (reflecting 1 unexpected sample swap and 2 participants with known sex chromosome aneuploidy), excessive homozygosity, or relatedness to another genotyped participant (pi-hat > 0.185). Due to high call rates, no individuals were excluded for excessive missingness, and variants were not excluded from analysis based on Hardy-Weinberg Equilibrium since the population is derived from a case-only study. For additional information, see the complete GWAS quality control protocol in the Supplemental Methods. A total of 281 unique individuals and 2,049,113 variants passed all quality control procedures.

Gene expression profiling was performed using the Illumina Human HT-12 Expression BeadChip for 255 study participants and generated in 4 batches. Quality control was performed using the limma R package ^53^. Samples with low signal-to-noise ratios or excessive proportions of undetected probes or systematically shifted expression levels (relative to other samples within the same batch) were excluded. All batches were background-corrected and quantile-normalized together using limma’s *neqc* function. Samples were excluded from analyses if they were flagged as a sample swap in analysis of each omics dataset or if they came from an individual that had an underlying genetic condition, as defined in the SOARS-B study^18, 50^. Probes were excluded if they displayed poor mapping quality or were not detected across all samples. Probes with a detection p-value ≥ 0.05 were set to missing for downstream analyses. For additional details, see the full protocol in the Supplemental Methods.

Methylation profiling was performed using the Illumina Infinium MethylationEPIC array for 277 study participants. Quality control and preprocessing were performed using a previously described methylation pipeline ^54^. In parallel, the R package ewastools ^55^ was used to examine sample-level Illumina quality control metrics and to confirm sample identity where possible. For additional information, see the complete EPIC array quality control protocol in the Supplemental Methods

### Candidate Gene Selection and Testing

Candidate genes were selected from canonical signaling pathways that interact with oxytocin directly or show strong patterns of regional co-expression within the brain (**Supplementary Methods, Targeted Genotyping**). Using this approach, the following KEGG-annotated pathways were chosen: “Oxytocin signaling pathway” (map0921); “Dopaminergic synapse” (map04728) ^56-58^; “Serotonergic synapse” (map04726) ^59, 60^; “Cholinergic synapse” (map04725) ^4, 61, 62^; “GABAergic synapse” (map04727); “Glutamatergic synapse” (map04724) ^63^; “Estrogen signaling pathway” (map04915). Genes from each KEGG pathway were extracted using the R package KEGGREST v 1.24.1 ^64^.

Redundant genes between KEGG pathways were removed, creating a final list of 494 genes. The UCSC table browser was used to retrieve GENCODE “knowncanonical” hg19 coordinates for each gene with a +/- 5 kb buffer to include proximal regulatory regions. The expanded coordinates were intersected with the “InfiniumOmni2-5-8v1-3_A1” manifest using BEDtools v 2.29.2 ^65^ to identify each probe within the candidate gene list. The resulting probe list was used in the targeted Omni 2.5 analysis.

TAQMAN genotyping candidates were identified using the UCSC table browser to extract the rsIDs of all common SNPs (build 138) that fell within the genomic coordinates (+/- 5 kb) of genes in the dopamine and oxytocin signaling pathways and retained if the rsID was not present in the Omni 2.5 manifest. Further filtering was performed by quantifying the number of published articles for each using the R package easyPubmed (v 2.13) with the following search terms: rsID AND (“plasma oxytocin” OR “serum oxytocin” OR “salivary oxytocin” OR “central oxytocin” OR “plasma OT “ OR “serum OT” OR “salivary OT” OR “central OT “) and choosing the most cited rsIDs from the results. A summary of these findings is provided in ^66^.

### Targeted Genotyping

Eleven TaqMan assays (Thermofisher, MA) (**Supplementary Table 1**) were selected based on the previously described literature review (**Supplementary Table 2**) in addition to a twelfth, which overlapped with the Omni 2.5 manifest on the X chromosome and was used for sample identity confirmation. Variant calling and quality assessments were performed using a ViiA 7 and the ViiA 7 software suite (ThermoFisher, MA), and sample-level quality controls were applied using PLINK (v1.9).

### Targeted OXTR Methylation Profiling

Targeted pyrosequencing was used to profile three CpG sites within the MT2 region of OXTR (hg19 - chr3:8,810,774-8,810,807) that were previously associated with ASD ^43^. Briefly, DNA from each participant was bisulfite converted, PCR amplified (in triplicate) and run on a Qiagen Q96 pyrosequencer to generate DNA methylation estimates for each CpG site. Samples were flagged and rerun if methylation values exhibited excessive variation (>5%) at one or more of the three CpG sites, or if they had fewer than two successful measurements for each CpG. Reported measurements are in units of mean percent methylation (across a minimum of 2 technical replicates for each CpG/sample). A detailed protocol including primer design, PCR programs, and analytical details is provided in the Supplemental Methods.

### CNV Analyses

CNVs were called for all samples genotyped on the Omni 2.5 array using PennCNV ^67^. After CNV and sample-level quality control procedures (Supplemental Methods), 259 participants displayed at least one autosomal CNV (>= 20kb in length) for a total of 1,198 CNVs. Frequency filtering was then applied to generate a dataset consisting of rare (5% frequency) CNVs for downstream analysis.

### Pathway Analyses

Pathway-level analyses were used to address our limited power to detect associations at the level of individual variants, probes, and CpGs in our molecular datasets. We profiled 5 pathway collections from the msigdb R package ^68^ across all three modalities: Hallmark, Gene Ontology (GO) Biological Process, GO Cellular Component, GO Molecular Function, and KEGG (supplemented to also include pathways selected of particular relevance to oxytocin, as described above). For our genomic dataset, we generated gene-level association p-values for rare and common variation present in autosomal genes utilizing the SKAT-O algorithm within the SKAT R package ^69^. We implemented an Over Representation Analysis using the 10% of genes with the lowest p-values (<= 0.053) from SKAT-O as ‘associations’ and tested against the background of all genes tested in SKAT-O (n = 24,501) present within each of the selected pathway collections. For the transcriptome and methylome datasets, we performed pathway-level analyses utilizing fast Gene Set Enrichment (fGSEA), as implemented in the fgsea R package ^70^. As input for these analyses, we leveraged individual probe/CpG results from our linear model association testing in limma to select a single probe/CpG per gene, based on lowest p-value for association with oxytocin, and then ranked genes based on the corresponding fold change for the oxytocin model term. Additional details can be found in the pathway analysis sections of the Supplemental Methods.

### Multi-Omic Analyses

Data from the DNA methylation, gene expression, and genotype arrays were used as inputs for the integrated analysis using the Multi-Factor Omics Analysis + (“MOFA+”) package in R ^71^. Approximately 8,300-8,500 genes per dataset were integrated to create 20 factors, which were then tested for association with plasma oxytocin. Details of the analysis are provided in the multi-omic sections of the Supplemental Methods.

### Statistical Analyses

All analyses were performed on plasma OT measurements obtained at screen or baseline study visits (i.e., pre-treatment), either as the dependent variable of interest or as the independent variable of interest (in transcriptome and methylome analyses executed using limma). We adjusted for a consistent set of covariates across all modalities that included functional strata as defined in the SOARS-B study ^18, 50^, age at sample, sex, 3 principal components reflecting genetic ancestry, and oxytocin measurement batch (which was correlated with participants’ study site). Modality-specific covariates were adjusted for as necessary and included gene-expression assay batch, OXTR assay plate, and a series of covariates to adjust for cell-type proportions and assay position within the methylome dataset. Linear model residuals, after accounting for covariates above, were used as input for the expression and methylation inputs for MOFA+, initially in univariate models and then in models controlling only for plasma OT measurement batch.

Association testing for all SNP-level genotypic analyses with plasma OT was conducted using PLINK 1.9. Association test results at the probe or CpG-level in our transcriptome and methylome datasets were only used for downstream pathway analyses because we were not adequately powered to detect signal from those datasets in this study.

CNVs were tested through burden tests implemented in CONCUR ^72^ and with generalized linear models in R, where burden was modeled as total number of CNVs and total length of CNVs in kilobases. All tests were performed within deletions and duplications combined, deletions-only, and duplications-only datasets.

## RESULTS

After the completion of quality control analysis (see Methods) at least one modality of omics data was available for 175 participants with most participants possessing data for analysis in all three modalities (Figure 1).

Of the overall cohort, participants were on average 10.7 years of age, 14 % were female, and 71% were white (Table 1). Among the individuals included in the analysis, the mean OT level was 2.3 ln pg/mL.

### Genome-Wide Common Variant Analysis

A total of 1,276,908 common (minor allele frequency (MAF) >= 0.05) autosomal and X-chromosome variants passed quality control and were tested for association with plasma oxytocin, adjusting for sex, functional strata, age at time of sampling, ancestry principal components, and oxytocin measurement batch (**Figure 2A**). These genome-wide analyses yielded a lambda of 1.008, suggesting adequate control of population stratification in our population (**Figure 2B**).

**Figure 2:**
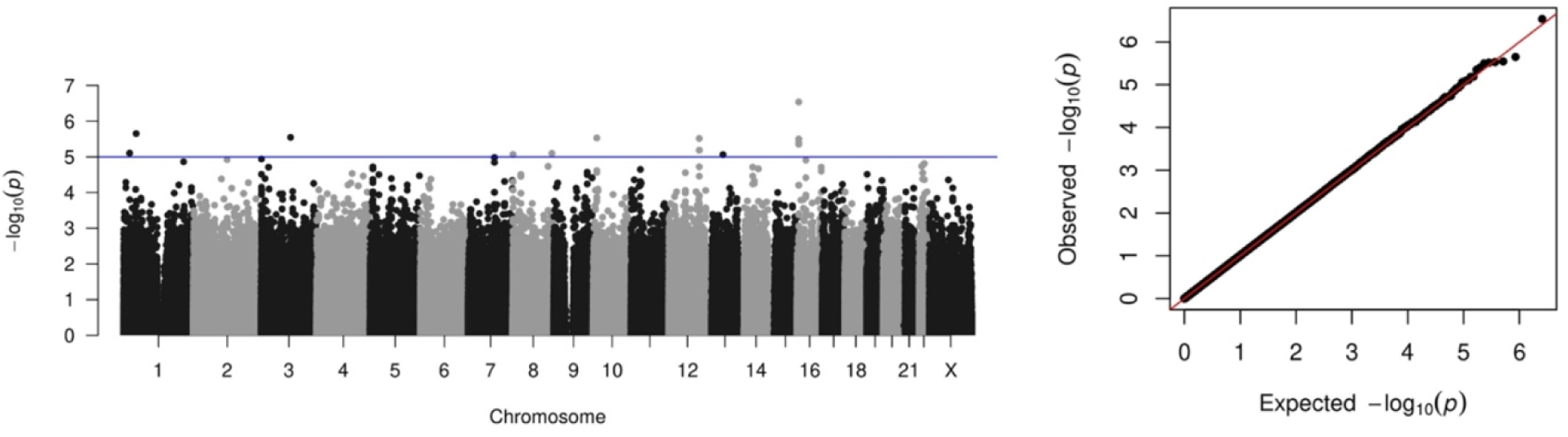
Genome-wide Common variant analysis of the SOARS-B cohort for association with plasma oxytocin. A) Manhattan plot showing 14 SNPs associated with plasma oxytocin (1 × 10^−5^) and B) Q-Q plot of our data showing appropriate control of population stratification.

Although no SNPs reached genome-wide significance, fourteen SNPs were associated with plasma OT at a threshold of 1 × 10^−5^, of which seven were intronic, four intergenic, two downstream of genes, and one in an annotated regulatory region (**Table 2**).

**Table 2:** Genome-wide suggestive SNP loci results from association with pre-treatment natural log-transformed plasma oxytocin.

| SNP | Chr (hg19) | Position (hg19) | Alleles | Beta (95% CI) | P-Value | Freq | Nearest Gene(s) |
| --- | --- | --- | --- | --- | --- | --- | --- |
| rs1780315 | 1 | 21863956 | G/A | -0.26 (-0.37 – -0.15) | 7.9E-06 | 0.27 | ALPL |
| rs12036829 | 1 | 45285664 | A/G | 0.56 (0.33 – 0.78) | 2.2E-06 | 0.06 | BTBD19, PTCH2, RNU5E-6P |
| rs62260734 | 3 | 104979397 | A/G | 0.36 (0.21 – 0.50) | 2.8E-06 | 0.13 | ALCAM |
| rs11785337 | 8 | 1169491 | A/G | 0.29 (0.17 – 0.42) | 8.5E-06 | 0.22 | DLGAP2 |
| rs7017524 | 8 | 139593796 | A/C | 0.31 (0.18 – 0.45) | 8.0E-06 | 0.16 | COL22A1 |
| rs75796442 | 10 | 13006376 | G/A | 0.43 (0.25 – 0.60) | 3.0E-06 | 0.09 | CCDC3 |
| rs803565 | 12 | 108786534 | G/C | 0.28 (0.17 – 0.39) | 3.0E-06 | 0.24 | CMKLR1 |
| rs2559883 | 12 | 108807177 | G/A | 0.31 (0.18 – 0.45) | 6.5E-06 | 0.15 | CMKLR1 |
| rs56208353 | 12 | 109297829 | A/G | 0.54 (0.32 – 0.77) | 6.5E-06 | 0.05 | DAO |
| rs7331972 | 13 | 60115787 | A/C | -0.64 (-0.92 – -0.37) | 8.6E-06 | 0.05 | RNU7-88P |
| rs9940013 | 16 | 6113392 | A/G | -0.41 (-0.58 – -0.24) | 3.2E-06 | 0.09 | RBFOX1 |
| rs6500746 | 16 | 6113709 | A/G | -0.33 (-0.45 – -0.21) | 2.9E-07 | 0.16 | RBFOX1 |
| rs8052564 | 16 | 6115382 | A/C | -0.40 (-0.56 – -0.23) | 4.5E-06 | 0.09 | RBFOX1 |
| rs11645768 | 16 | 6120149 | G/A | -0.40 (-0.57 – -0.24) | 4.0E-06 | 0.09 | RBFOX1 |

Alleles displayed as effect allele/reference allele
CI is confidence interval.
Beta, CI & P-Value report on the additive genetic model effects, adjusted for sex, functional strata, ancestry principal components, and oxytocin measurement batch.
Freq is the allele frequency of the effect allele in the analysis population.

The most significant polymorphism, rs6500746, falls within the first intron of the RNA binding Fox-1 Homolog 1 gene (*RBFOX1*), and had a negative association with plasma OT levels (β = − 0.33, p-value 2.9 × 10^−7^) (**Supplementary Figure 2**). Three additional common *RBFOX1* variants exhibited similar directions of effect at a suggestive significance threshold (rs9940013, rs8052564, and rs11645768) (**Figure 3**). These four variants fall within a 7 kb region of the large first intron of *RBFOX1* and are included in three haplotypes commonly found in individuals with European ancestry as reported by LDlink ^73^. Conversely, a SNP in the first intron of *DLGAP2*, a gene which has previously been associated with ASD ^74^, displayed a positive association with plasma OT in our study population (rs11785337, β = 0.29, p-value 8.5 × 10^−6^).

**Figure 3:**
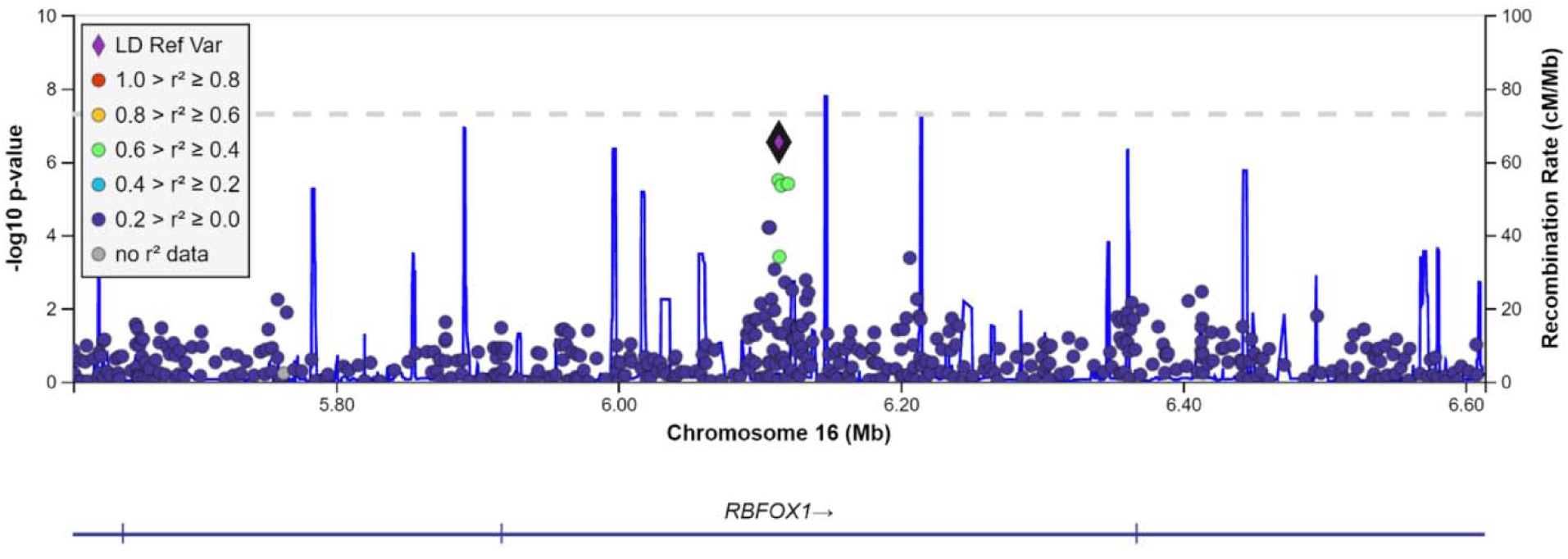
Locus Zoom plot for RBFOX1, the most significantly associated region with plasma oxytocin levels using a common SNP approach.

### Genotype Analyses within Pre-Defined Genes of Interest

Variants within genic regions defined by pathways of interest (Supplemental Methods) were filtered for those with at least three minor alleles observed in our analysis population and tested for association with plasma oxytocin. Interestingly, all the six variants with a suggestive association with plasma OT were carried by individuals whose ancestry was self-reported as African (n = 3), no self-reported ancestry (n = 1) or mixed (n = 1) ancestry (**Table 3**).

**Table 3:** Genome-wide suggestive SNP loci (within genes of interest) results from association with pre-treatment natural log-transformed plasma oxytocin.

| SNP | Chr (hg19) | Position (hg19) | Alleles | Beta (95% CI) | P-Value | Freq | Gene |
| --- | --- | --- | --- | --- | --- | --- | --- |
| rs3842724 | 11 | 2185556 | A/G | 1.14 (0.66 – 1.61) | 5.0E-06 | 0.011 | TH |
| rs17124695 | 14 | 52339300 | G/A | 1.27 (0.74 – 1.80) | 6.2E-06 | 0.009 | GNG2 |
| rs17124713 | 14 | 52349410 | A/G | 1.27 (0.74 – 1.80) | 6.2E-06 | 0.009 | GNG2 |
| rs17092983 | 14 | 52357476 | A/G | 1.27 (0.74 – 1.80) | 6.2E-06 | 0.009 | GNG2 |
| rs7342769 | 16 | 23845262 | G/A | 1.27 (0.74 – 1.80) | 6.2E-06 | 0.009 | PRKCB |
| rs7186490 | 16 | 24288512 | G/A | 1.05 (0.61 – 1.49) | 5.9E-06 | 0.014 | CACNG3 |

The most significant SNP from this analysis, rs3842724, falls within exon 14 of the Tyrosine Hydroxylase (*TH*) gene and is associated with a relatively larger increase in plasma OT levels (β = 1.14, p-value 4.98 × 10^−6^). The ‘A’ effect allele has an AFLA-estimated ^75^ global MAF of ∼0.008, with higher representation in African and African American genomes (0.07 - 0.09). Three additional suggestive variants, rs17124695, rs17124713, and rs17092983, fall within a 18,176 bp region of G Protein Sub-Unit Gamma 2 (*GNG2*). This apparent haplotype is present in ∼5.8% of individuals with African ancestry as reported in LDLink ^73^ and is associated with a β-value of 1.27 in the three heterozygous carriers of the G, A, A minor alleles, respectively.

Another significant SNP, rs7186490 (β = 1.05, p-value 5.9 × 10^−6^), falls in the first intron of the Voltage-Dependent Calcium Channel Gamma-3 subunit (*CACNG3*). Like the other top variants, the AFLA-estimated frequency of the minor A-allele of rs7186490 is virtually absent in European genomes (0%) and more common in African and African American genomes (∼9%). Our cohort contained five heterozygous carriers of the A-allele with a study MAF of 0.014, making it one of our more common variants among the suggestive polymorphisms from the targeted analysis.

The variant rs7342769 is located ∼2,000 bp upstream of Protein Kinase C Beta (*PRKCB*) and associated with increased levels of plasma OT (β = 1.27, p-value 6.2 × 10^−6^, study MAF = 0.009). ALFA estimates show a low global prevalence of the G allele (MAF = 0.0048), and it is extremely rare in European populations (MAF = 0.000384). However, it has a relatively higher prevalence in African / African American populations (MAF = 0.075 - 0.089), and all G-allele carriers in this study are of self-report African or African American ancestry.

Several associations were statistically less significant but exhibited much higher MAFs within our study population. These include variants in *GABBR1* (rs926552, MAF = 0.12, β = 0.3151, p-value <10^−3^, Global MAF=0.10) and *ADCY2* (rs326153, MAF = 0.055, β = 0.54, p-value <10^−3^, Global MAF=0.04), which exhibit relatively high population frequencies in both our study and in the general population.

The R package SKAT^76^ was used to examine the association of rare variants aggregated at the gene level with plasma OT by including rare variants (those with a minor allele count of 1 to those with a minor allele frequency of 1%) that passed quality control and were present within our genes of interest. Three genes (*ESR1, GNG2*, and *PLCL1*) passed a family-wise error threshold of 0.1 and had nominal p-values ranging from 1.1 × 10^−4^ to 3.0 × 10^−4^ (**Table 4**) akin to data from the *GNG2* region (**Supplementary Table 8**).

**Table 4:**
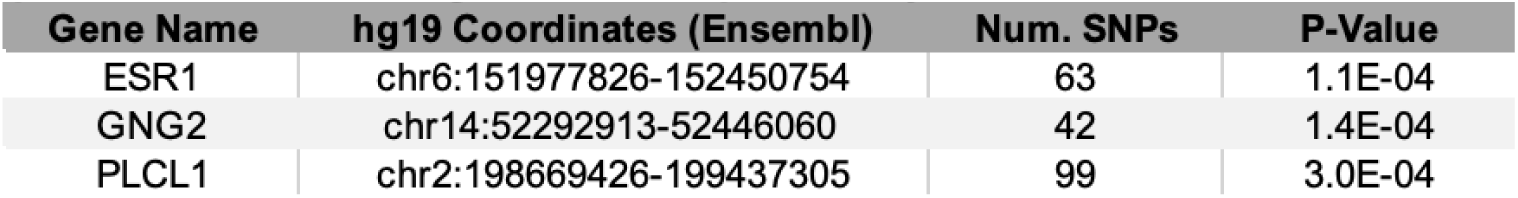
Candidate gene SKAT results (FWER<0.1) from association with pre-treatment natural log-transformed plasma oxytocin.

| Gene Name | hg19 Coordinates (Ensembl) | Num. SNPs | P-Value |
| --- | --- | --- | --- |
| ESR1 | chr6:151977826-152450754 | 63 | 1.1E-04 |
| GNG2 | chr14:52292913-52446060 | 42 | 1.4E-04 |
| PLCL1 | chr2:198669426-199437305 | 99 | 3.0E-04 |

### Targeted Variant Analyses

None of the eleven targeted genotype assays were significantly associated with plasma OT levels, failing to replicate the findings of previous studies (**Supplementary Table 3**).

### Association of Methylation at OXTR Locus

In analyses examining the relationship between pre-treatment plasma OT levels and methylation at the OXTR locus, 172 participants had complete data available after quality control. There was no significant association between plasma OT levels and the three CpGs individually (−924, p = 0.11 & −934, p = 0.41), as well as the mean methylation of all three CpGs (p = 0.47) (**Supplementary Table 4**) with the design described in **Supplementary Table 5**.

### CNV Burden Testing Analyses

Rare CNV burden was not significantly associated with plasma OT in any CONCUR or basic linear model analysis (**Supplementary Tables 6** and **7**). Furthermore, among the generalized linear model analyses, only rare deletion and duplication CNV burden, when modeled as total number of CNVs, had a nominal negative association with pre-treatment plasma OT (β = −0.0005, p-value 0.03).

### Genomic, Transcriptome and Methylome Pathway Analyses

Significant associations (FDR-BH < 0.10) were detected across all the included gene sets (Gene Ontology terms: Biological Process, Cellular Component, Molecular Function; MSigDB – Hallmark, MSigDB – KEGG). The top five significant associations (when available) for each pathway are presented in (**Table 5 and 6**) and unfiltered results with the leading-edge genes are provided in (**Supplementary Data 1**). There was very little overlap between the leading edges of the significant transcriptomic and DNA-methylation gene sets (**Supplementary Data 2**) indicating that these signatures were unlikely to represent direct interactions between DNA methylation and transcriptional activity.

**Table 5:**
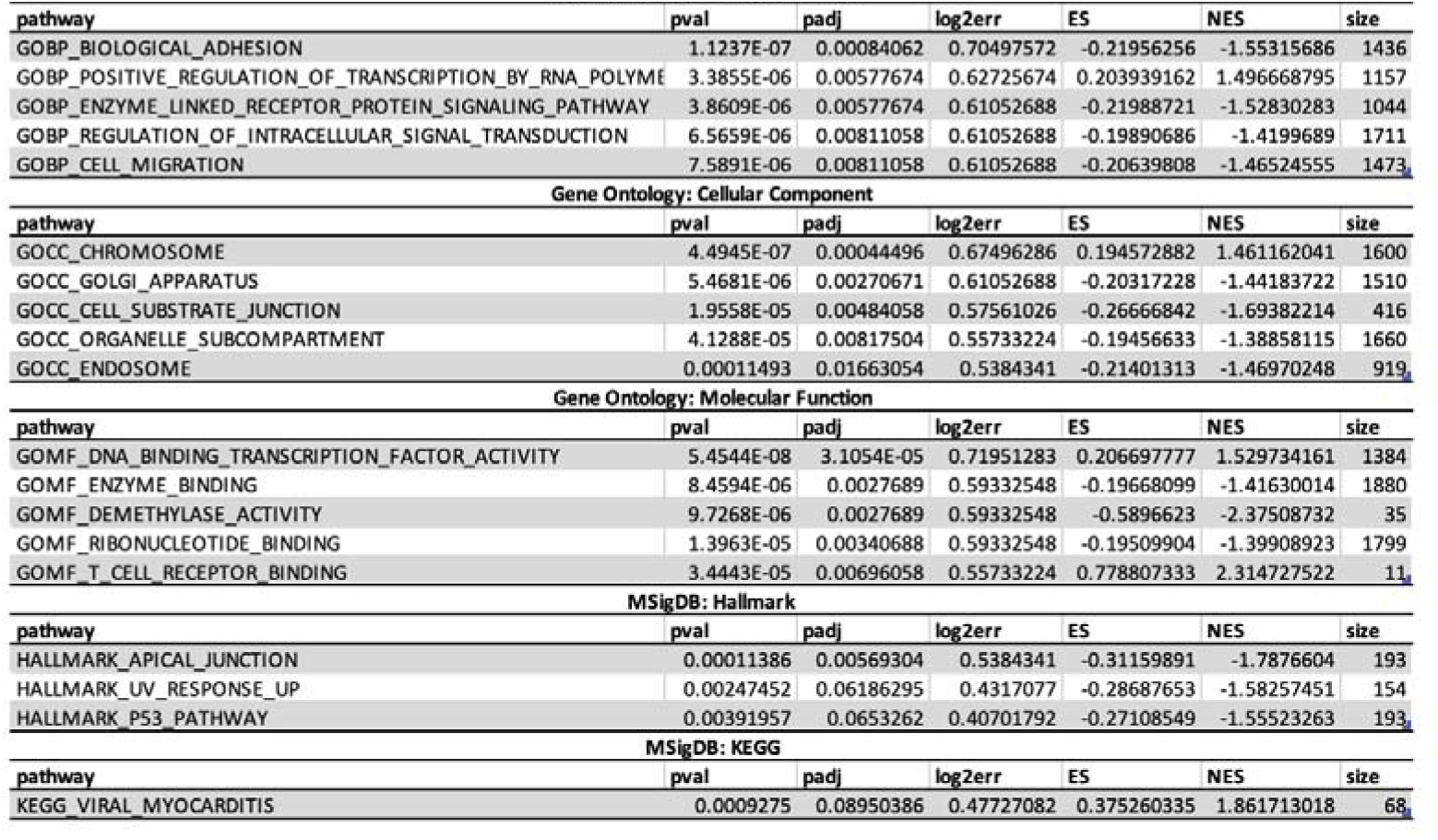
Differential DNA methylation: Top 5 Gene-Set Enrichment Results Gene Ontology: Biological Process.

**Table 6:**
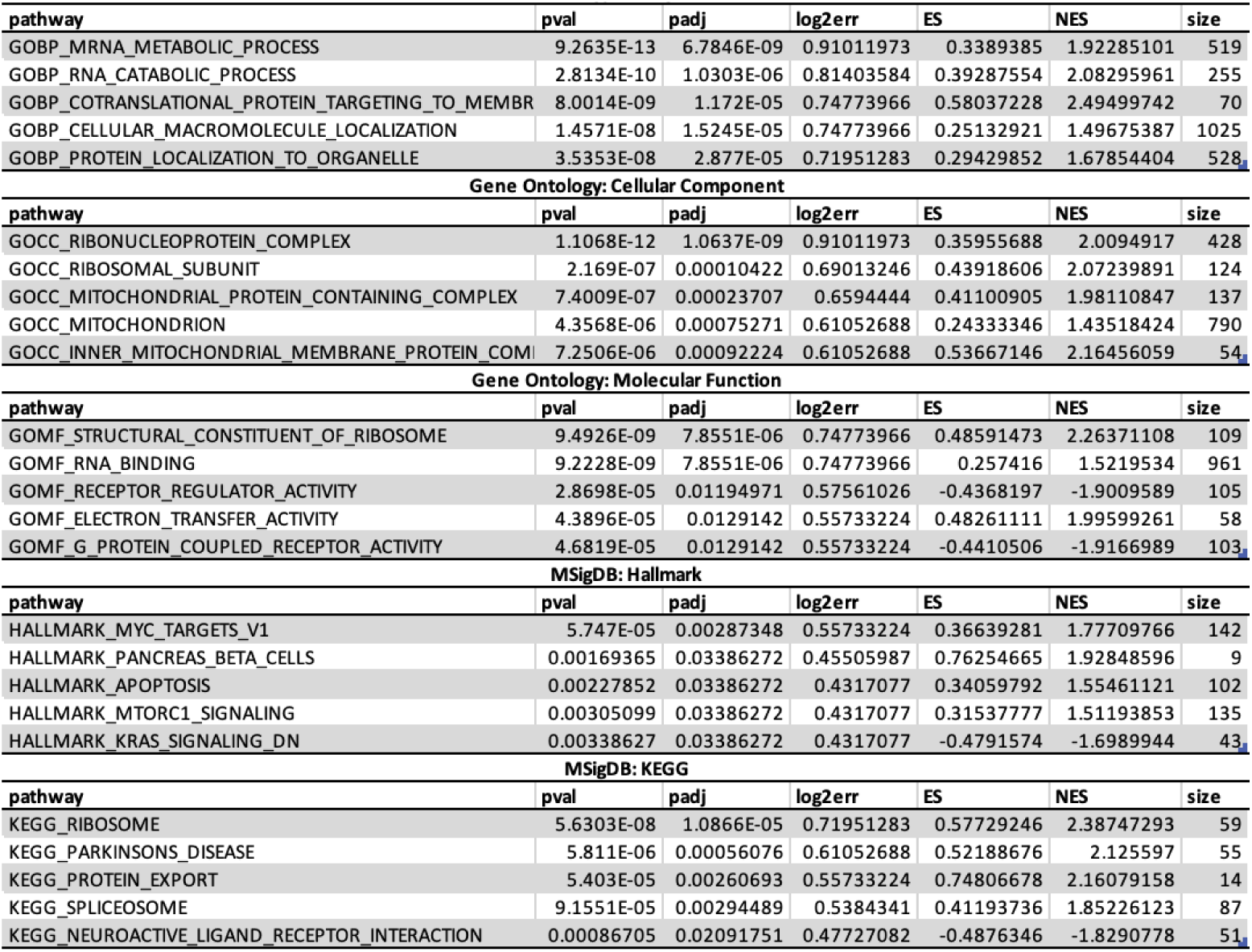
Differential Gene Expression: Top 5 Gene-Set Enrichment Results Gene Ontology: Biological Process.

### Multi-Omics Analyses

The integrated omic analysis did not reveal additional correlation among the individual omic platforms. The resulting MOFA+ factors were each dominated by one input dataset, with the gene expression dataset having the predominant variance explained over all factors and the genotype dataset having the lowest (<10%; data not shown). In univariate models, factors 6, 8, 12 & 17 met an FDR-adjusted p-value < 0.10, but after testing all factors in models adjusting for oxytocin measurement batch, factors 8 (β=0.30, p-value=0.009), 11 (β=0.06, p-value=0.026), and 16 (β=0.09, p-value=0.040) were nominally significant with positive associations at an unadjusted p-value of 0.05 but not after FDR adjustment (**Supplementary Table 9**). These nominally significant factors represented the individual inputs of methylation (factors 8 and 16) and gene expression (factor 11) datasets.

## DISCUSSION

Our study represents the most comprehensive genetic, epigenetic, and transcriptomic analysis of endogenous plasma OT levels to date. Our findings provide important insights into the genetic underpinnings of oxytocin signaling and the peripheral correlates of circulating oxytocin with gene expression and DNA methylation. They help to address a paucity of currently available data about the molecular regulation of endogenous oxytocin in humans. Targeted genotype analyses identified several variants located in genes from the oxytocin signaling pathway and other closely related pathways, and additional candidates were identified at a suggestive threshold in the unbiased genome-wide analysis. Some of these variants overlap with annotated ASD risk genes and show functional enrichment in Ca^2+^ related activity in dopamine and oxytocin signaling pathways. The associations between these variants and plasma OT levels in humans, albeit suggestive, are novel and substantially broaden the number of candidate genes that are now associated with endogenous oxytocin levels. In contrast, gene expression and DNA methylation results did not identify any individual genes or methylation markers that were associated with plasma oxytocin. However, both datasets did show intriguing patterns of pathway enrichment that could indicate a role for plasma OT in modulating a wide array of biological processes in the periphery, particularly in the domains of cell adhesion, immune activity, and cell-to-cell signaling.

### Genotype Results

Unlike the peripheral methylation and expression data, genotypes that are associated with plasma OT levels could indicate neurologically associated mechanisms. Prior to our study, only a small number of variants had been suggestively associated with plasma OT levels. Two previous studies identified a borderline association between rs2254298, which is in the third intron of *OXTR*, and plasma OT levels in individuals with ASD ^77, 78^, but these were not significant after correction for multiple testing. A third study identified an association with rs12625893, which is located ∼20 kb downstream of *OXT*, in a study cohort with obesity and impaired glucose tolerance ^49^. We did not replicate either of these prior findings but do identify new suggestive associations for plasma OT as a quantitative trait.

The SOARS-B study, from which our samples were derived ^18^, allowed us to include non-Caucasian participants to identify suggestive genetic associations that might be unique to specific ancestries. The benefit of this approach is reflected in multiple suggestive associations that were only present in individuals of self-report African ancestry. The top hit in this category is for the A allele of rs3842724 in tyrosine hydroxylase (*TH*), which is associated with a large increase of plasma OT in A-allele carriers (β = 1.14). This is of particular interest because TH is responsible for catalyzing L-tyrosine into L-3,4-dihydroxyphenylalanine and is the rate-limiting enzyme in the biosynthesis of catecholamines, including dopamine ^79^. In humans, approximately 40% of neuronal cells in the SON and PVN of the hypothalamus show positive immunostaining for TH, and there is substantial overlap between TH+ and OXT+ magnocellular neurons, which are the primary source for oxytocin release in the brain ^80^. In rats, activation of dopamine receptors in the PVN stimulates the release of oxytocin and impacts the activity of mesolimbic dopaminergic neurons ^81^. Changes in TH levels or functionality in these neurons may impact the autoregulatory positive feedback loop between dopamine and oxytocin release ^82^ and contribute to the increased levels of plasma OT observed in the A-allele carriers in our study.

We also identified increased plasma OT in C-allele carriers of rs7186490, which is in the Calcium Voltage-Gated Channel Auxiliary Sub-Unit Gamma 3 (*CACNG3*) gene. CACNG3 is a component of one of the calcium channels that opens in response to OXTR activation. This leads to depolarization and the subsequent release of oxytocin and other neuropeptides from the activated neuron. This is an important mechanism for regulating activity in the hypothalamic-neurohypophysial system, which is heavily influenced by voltage-gated Ca^2+^ channel density ^83^ and impacts the release of oxytocin ^84^; however, there is very little published data on the role of *CACNG3* in the brain. Two recent studies implicated *CACNG3* in addiction ^85^ and memory deficits ^86^ in the hippocampus, both of which have also been associated with disruptions to oxytocin signaling. These findings point towards a potential role for CACNG3 in the activation of oxytocin-associated neurons, its broader overlap with glutamatergic signaling, and modulation of OT release via Ca^2+^ depolarization, which might contribute to the increased plasma OT levels observed in the five C-allele carriers of rs7186490.

There is less autism related literature for G Protein Subunit Gamma 2 (*GNG2*), which was associated with a similarly large increase (β = 1.27) in 3 heterozygous carriers of the “GAA” haplotype across variants rs17092983, rs17124695, and rs17124713 ^87, 88^. *GNG2* is predicted to downregulate both Ca^2+^ channel activity and adenylate cyclase activity while upregulating phospholipase C activity, which is one of the first downstream targets of *OXTR*. Interestingly, the three carriers of the GNG2 GAA haplotype were also carriers of the G allele of rs7342769 in *PRKCB*, a direct downstream target of *OXTR* that modulates multiple secondary pathways including MAPK, Ras, and calcium ion signaling, with broad activity in the brain through the phosphorylation of target proteins ^89^. These two genes are on separate chromosomes, so the overlap of these three individuals is not based on linkage disequilibrium but could be driven by genetic background. Notably, *PRKCB* is also a strong candidate for ASD, with several studies supporting its association ^90-92^.

While all suggestive associations in the targeted analyses were centered around a relatively small number of individuals of self-reported African or multiracial ancestry, there was a borderline candidate allele with a much higher frequency within our cohort. The A-allele of rs926552 in the Gamma-Aminobutyric Acid Type B Receptor Subunit 1 gene (*GABBR1*) was represented in our cohort by one homozygous carrier and 40 heterozygous carriers (study MAF = 0.12 vs. ALFA estimated global MAF = 0.1) and associated with a moderate increase in plasma OT (β = 0.31, p = 1.58 × 10^−4^).^92-94^ *GABBR1* expression levels ^95, 96^ and methylation levels ^97^ are also significantly altered in the brains of individuals with ASD.

Among the results of the genome-wide analysis, the association between the A-allele carriers of rs6500746 in the RNA Binding Fox-1 Homolog 1 gene (*RBFOX1*) and reduced plasma OT (β = − 0.33) is the most robust. Functionally, *RBFOX1* encodes an RNA-binding protein that regulates tissue specific nuclear RNA splicing ^98-100^ and enhances cytoplasmic mRNA stability ^101^, however it is also strongly associated with ASD ^102^. Knockout of *Rbfox1 in vitro* and using *in vivo* mouse models results in abnormal brain development, with deficits in radial migration of cortical neurons, dendritic arborization, and electrophysiological characteristics ^103, 104^. At the time of writing, there are no published connections between *RBFOX1* and oxytocin signaling or plasma OT in the literature. However, a recent study shows that RBFOX1 regulates synaptic transmission and excitatory-inhibitory (E/I) balance ^105^, which could have a direct impact on the release of oxytocin based on activity-dependent release.

Finally, we established that the A-allele of rs11785337 was associated with a moderate increase in plasma OT (β = 0.29, study MAF = 0.22). The SNP is located within the DLG Associated Protein 2 gene (*DLGAP2*) that has a suggestive link to ASD ^106, 107^. The *DLGAP2* encodes a membrane-associated protein that serves as a major a structural component (along with DLGAP1, 3 and 4) of the post-synaptic density, where it impacts glutamatergic signaling activity ^108^. A CRISPR-based silencing of *DLGAP2* in an *in vitro* model of iPSC-derived excitatory neurons revealed reduced spontaneous excitatory postsynaptic currents, which further implicates the gene in maintaining E/I balance ^109^. Signatures of this disruption are also present in a mouse *Dlgap2*(-/-) model of ASD, in which double-knockout mice exhibit increased aggressive and dominance-related behavior, altered dendritic spine density, decreased peak amplitude in excitatory post-synaptic current, and reduced post-synaptic density ^110^.

The gene level analyses of rare variants further supported the association of plasma OT with *GNG2* and identified an association with Estrogen Receptor 1 (*ESR1*) and Phospholipase C-like 1 (*PLCL1*). *ESR1* was previously shown to regulate OT signaling during gestation and parturition ^111, 112^ and variation in *ESR1* is associated with social interaction and emotional regulation in individuals with ASD ^113^, but it has not been previously associated with circulating OT levels. Similarly, *PLCL1* has been associated with ASD ^114^ and plays an inhibitory role in uterine contraction through modulation of the OT signaling pathway ^115^, but has not been reported in modulating plasma OT levels. These findings build upon a growing body of evidence for the roll of these genes in OT signaling and social behavior.

### Gene Expression and DNA Methylation

#### Gene Expression

The gene set enrichment analysis (GSEA) of changes in peripheral gene expression associated with differences in plasma OT identified several immune-related pathways, including “immune response”, “immune activation”, “protein tracking”, and “catabolic processes/oxidative phosphorylation”. These findings corroborate a growing body of research that suggest an immunoregulatory role of oxytocin in both CNS and peripheral contexts ^116^, and a functional and mechanistic coupling between the immune and neuroendocrine system more broadly ^117^. Cytokines like IL-1, IL-2, IL-6, interferon-gamma, and transforming growth factor-beta are produced by hypothalamic cells ^118^, which may play a role in mediating the production and release of neuropeptides ^119^.

All but 1 of the 26 significant GO:BP associations(adjusted p value <0.1, see **Supplementary Data 2**) were positively associated with plasma OT levels. This overarching trend of increased transcriptional activity with plasma OT could indicate a relationship between neuronal oxytocin production and enhanced immune system activity that has been reported in ASD ^120-122^. This type of interaction has been observed more directly using *in vitro* models, such as uterine smooth muscle cells, which increase the expression of *OXTR* following exposure to IL-6 ^123^, or in cultured microglia, which increase *OXTR* expression in response to bacterial endotoxin and show reduced activation when pre-treated with endogenous oxytocin ^124^.

G protein coupled receptor activity was also enriched in the GO and molecular function annotations and negatively associated with plasma oxytocin. These findings may also overlap at a functional level with the broader implication of immune system activity with plasma OT levels. Immune cells rely heavily on GPCR signaling to navigate ^125^ and regulate context-specific inflammatory processes ^126^. Terms related to secretion were positively enriched, including “secretory granule”, “vesicle”, and “vesicle lumen”, which have functional roles in releasing neuropeptides and hormones from the neuroendocrine system and effector molecules in the peripheral immune system.

### Methylation Results

#### Genome-Wide Methylation

DNA methylation is generally considered to be a more stable molecular measure than gene expression ^127^. Based upon prior reports, correlations between blood and brain methylation markers appear to be poor, with one study showing ∼7% of CpGs significantly correlated between blood and brain tissue ^128^. Even so, peripheral markers that are not directly correlated with brain methylation could still be useful if they are altered due to neurological phenotypes that can affect the periphery, such as variation in plasma OT levels. In the case of *OXTR*, positions −901, −924, and −934 were we previously reported to be hypermethylated in both peripheral blood mononuclear cells and post-mortem cortical samples of individuals with ASD, lending to the idea that early developmental insults could impact *OXTR* methylation prior to establishing the primordial germ layers ^43^, and that those changes in methylation are maintained into adulthood.

As was the case in the gene expression analysis, we did not find any probe-level (CpG resolution) methylation associations with plasma OT after correcting for multiple testing, nor did we find any associations in the aggregated regional analysis. However, GSEA identified several enriched terms. Within the Hallmark gene set, genes involved in apical junctions were negatively enriched, which is consistent with the broader theme of cell adhesion terms that were present in the gene expression analysis. Of the 46 GO biological pathway results, there was positive enrichment of DNA methylation with plasma OT in 13 pathways that was largely associated with early embryonic / organ-level development and morphogenesis. Among the negatively enriched pathways, there are several terms that point to neuronally associated processes (i.e., axon development, receptor localization to synapse), which implies that, in aggregate, peripheral hypermethylation in these pathways may be associated with lower levels of plasma oxytocin. Interestingly, “demethylase activity” was the top negatively enriched gene set in the GO molecular function analysis and included several *KDM*-family genes. These genes are lysine-specific histone demethylases that broadly regulate gene expression and DNA repair. Deletions and splice variants of KDM5-family enzymes have recently been associated with intellectual disability and mild ASD ^129^. Finally, GO cellular component results largely aligned with the significant pathways identified in the biological pathway and molecular function analyses. As was the case in the biological pathway results, we once again see negative enrichment in terms like “axon”, reflecting a potential connection to neuronally relevant mechanisms in the brain.

### Targeted OXTR Methylation

The initial findings by our group ^43^ generated significant interest around the three differentially methylated sites in the promoter region of *OXTR* (−901, −924, −934) and subsequent findings continue to suggest that this region has a role in regulating oxytocin signaling in the brain. For instance, methylation levels between these sites are tightly correlated and have consistently been implicated in regulating *OXTR* expression ^130^. Furthermore, a recent study suggested that hypermethylation in −934 and −924 moderated the way that early life stress affects brain activity when anticipating rewards ^131^. However, findings in ASD remain inconclusive ^132^. Reports of associations with plasma OT levels have been inconsistent, though trends indicate that there is likely no association of endogenous oxytocin levels with methylation at these sites ^133, 134^, and our data support this lack of association.

### LIMITATIONS

This study had several limitations. First, the 290-person study cohort, while the largest to date, was insufficiently powered for genome-wide statistical analyses and further complicated by the diverse array of ages, ancestries, and the inclusion of both sexes. As a result, we may have missed small or moderate effect size associations with plasma OT levels, particularly for probe-level analyses, and our suggestive findings should be interpreted accordingly. We also acknowledge the limitations of using peripheral blood samples to identify biomarkers for OT levels, particularly in the gene expression and DNA methylation analyses. We chose this approach because it enabled us to perform the molecular analyses in a responsive tissue ^135^ that was collected during the same blood draw. Furthermore, several studies in separate fields have shown peripheral expression and DNA methylation profiles can be informative of states in the brain ^136, 137^. Finally, we acknowledge that peripheral OT has a half-life of less than an hour and may not be adequately represented by a single time point sample, compounded by the inherent variability in measuring endogenous OT using available assays ^66, 138, 139^.

## CONCLUSIONS

Our genetic analyses identified several novel associations between plasma OT and variation in *RBFOX1, DLGAP2, TH, GNG2, ESR1*, and *PLCL1*. Many of these genes have been previously associated with ASD, but their interactions with oxytocin signaling are not well characterized. They represent a cross-section of biological roles that include Ca^2+^ depolarization, G protein / neuropeptide signaling, RNA-metabolism, and phospholipid cleavage all of which can be connected to oxytocin signaling or its downstream mechanisms. However, additional studies will be required to validate these findings, to understand the mechanisms through which they impact circulating OT levels. Our gene expression analyses did not identify significant associations between plasma OT and gene expression at the level of individual probes or genes. However, GSEA identified associations with broader patterns of pathway-level changes including immune function, mRNA metabolism, cell adhesion, and G protein signaling, among others. Similarly, no significant associations were found between individual DNA-methylation sites and plasma OT, but GSEA identified several significant terms that broadly overlap with those identified in the expression data, though with little overlap in the genes driving these associations. Collectively, these novel associations between genetic variants and biological pathways with plasma OT levels provide exciting new candidates for future studies and add to the growing body of knowledge related to the complex etiology of ASD and its interactions with OT signaling.

## Data Availability

All data produced in the present study are available upon reasonable request to the corresponding author

## ACKNOWLEDGMENTS

First and foremost, we would like to thank the participants of the SOARS-B clinical trial and their caretakers who made this research possible. We would also like to thank the clinical sites from Duke University, the University of North Carolina Chapel Hill, Vanderbilt University, Massachusetts General Hospital, the Mount Sinai School of Medicine, and Seattle Children’s Hospital Research Institute that performed the recruitment, behavioral evaluations, and sample collections for the SOARS-B participants. Sheryl Walker, Ph.D., generated the plasma OT measurements. We would also like to thank David Layfield and the staff at Duke Biofluids Shared Resource, Dr. Devos and the rest of the staff at Duke’s Sequencing and Genomics Technologies Core, and the staff at the Duke Molecular Genomics core, particularly Karen Abramson, Stephanie Arvai, and Emily Hocke for their excellent support throughout the course of this study.

