## Supplementary Methods for "Genetic and epigenetic signatures associated with plasma oxytocin levels in children and adolescents with autism spectrum disorder"

**Supplemental Information**

### Candidate Gene Selection

#### Genome-Wide

Pathways and genes were selected for the targeted analysis from the KEGG-annotated canonical pathway for oxytocin signaling and from other pathways known to interact with oxytocin: "Oxytocin signaling pathway - Homo sapiens" (hsa04921); “Estrogen signaling pathway” (map04915) (CITE); "Dopaminergic synapse" (map04728) (1-3); "Serotonergic synapse" (map04726) (4, 5); "Cholinergic synapse" (map04725) (6-8); "GABAergic synapse" (map04727) / "glutamatergic synapse" (map04724) (9). Genes from each of these KEGG pathways were extracted using the R package KEGGREST (v 1.24.1 (10). An additional non-redundant gene list was selected from the literature that represented updated publications since the last published KEGG entry, including additional genes for dopamine (n = 9), arginine vasopressin (n = 1), acetylcholine (n = 3), GABA (n = 10), and glutamate (n = 20).

In addition to the updates in KEGG, an additional review of the literature was conducted to identify immune-related genes that are impacted by oxytocin, which are likely differential gene expression candidates in the blood. There are several studies evaluating the immunomodulatory effects of oxytocin including its role in ischemic injury, cardiovascular health, parturition, and in regulating peripheral and central immune cell activity. Using the search terms ("oxytocin" AND (immun* OR inflammation OR cytokine)) returned several publications that identified several immune-related genes that were included in the targeted analysis. These include *TNF-alpha*, *IL1RN*, *IL-4, IL-6*, *CCL3*, *CCL4*, *MCP-1*, *CXCL10*, and *VEGF* from a study that found exogenous oxytocin attenuated the inflammatory response to endotoxin stimulation in male human plasma samples (11); *IL-1β*, *COX-2*, *iNOS*, *MAPK1*, and *MAPK14* from a study that showed oxytocin attenuated cultured mouse microglial activation and cytokine release in (12); and finally. *CXCL8*, *CCL5*, *CCL2*, *SOD2*, *KAT5*, *PTGES2*, *PTGES*, *PRKCZ*, *ARRB2*, *ARRB1*, *KCNMA1*, and *TRPV2*, which were identified as having an immunoregulatory and other socially / mechanistically relevant functions in oxytocin (13).

Using the KEGGREST and biomaRt packages within R (14), hg19 coordinates were obtained for a total of 494 genes and +/-5000 bp were added to either side of their gene coordinates to include proximal regulatory regions. These resulting gene regions were used in downstream genotype-based analyses.

#### Targeted Genotyping

A number of genes have previously been associated with oxytocin function but may not be captured on the Illumina Omni 2.5 array. We first extracted rsIDs of all common snps (build 138) that fell within gene-based coordinates of genes within the dopamine and oxytocin signaling pathways that were not already present on the genotyping array (**Supplementary Figure 3**). These rsIDs were then evaluated using a PubMed scraper built around the R package easyPubmed (v 2.13). The SNPs were then prioritized based literature hits for each rsID that were associated with peripheral oxytocin and then filtered by European ancestry MAF threshold of >= 5% from dbSNP. Candidates SNPs were further screened using the NIH LitVar tool (15), which provides detailed research summaries of each variant. This method resulted in 2 candidates in *OXTR* (rs53576, rs2254298) and 2 candidates in *CD38* (rs3796863, rs6449182) for association with peripheral oxytocin levels. Additional SNPs were selected from genes directly involved in the production of oxytocin (*OXT*) or its Ca^2+^ induced release (*CACNA1C*) which have been previously associated with schizophrenia, autism, bipolar disorder, and/or depression; also filtered by a European MAF >= 5% and investigated in LitVar (**Supplementary Table 1**).

#### OXTR Methylation

Based on the findings of Gregory *et al.* (16) that identified association between hypermethylation of several CpGs in the promoter region of *OXTR* with ASD in both peripheral blood and post-mortem temporal cortex samples and because these sites are not captured on the Illumina EPIC methylation array, we decided to specifically assay these sites.

### Sample Collection and Processing

Blood, saliva, and plasma samples were obtained at the respective clinical sites (17) and shipped to the Duke Biofluids Shared Resource for extraction using an Autopure platform with Puregene chemistry (Qiagen, MD) (<9/2016) and a ReliaPrep™ Large Volume HT gDNA Isolation System on a HSM2.0 instrument (Promega, WI) (>9/2016). RNA samples were extracted using the QIAsymphony PAXgene Blood RNA Kit (Qiagen, MD).

### Plasma Oxytocin Quantification

A total of 7 mL of blood was drawn into lavender-top tubes and centrifuged to extract plasma. Plasma oxytocin levels were quantified in each sample using the Enzo Oxytocin ELISA kit (CAT: ADI-901-153A) with the standard protocol in 96-well batches at University of North Carolina, as previously described (18). Each sample was quantified using a standard curve on a plate reader.

### Genome-Wide Genotyping

Samples were randomized into batch units (8-sample Omni 2.5 arrays) to mitigate batch effects by ensuring that no array chip was over-enriched for sex, functional strata, or randomized treatment group. One sample from each individual was selected for the analysis based on their total available quantity (ng) and DNA integrity scores. Two standard Illumina protocols were used to prepare 200 ng of DNA from each individual and hybridize it to the array - the Infinium LCG Assay Manual Workflow (15023140 Rev. A, March 2011), which was followed up to the point of transferring the prepared samples onto the arrays for hybridization, and the Infinium LCG Assay Automated Workflow (15023141 Rev. A, March 2011), which was followed to complete the protocol prior to imaging the chips on the iScan.

### Targeted Genotyping

On-demand Taqman probes were purchased from Thermo Fisher (Waltham, MA) (**Supplementary Table 1)**. Genotyping reactions were prepared for 286 study participants for each of the twelve target variants and an X-linked control variant (rs17148391, assay ID: C_33857500_10) that was used to confirm genotype agreement with participants’ Omni 2.5 genotypes at that SNP.

### Transcriptome Profiling

Participants with a viable RNA sample from baseline or screen (> 0.05ug) and at least one other study visit were selected for measuring gene expression using the Illumina Human HT-12 gene expression array. Keeping all samples attributed to a participant together on arrays, participants were randomized into batch units (12-sample arrays) to minimize over-enrichment of sex, functional strata, or randomized treatment groups across each array. RNA samples were thawed on ice after being stored at -80°C and aliquots were normalized to 10 ng/µL in nuclease-free molecular-grade water. For each sample, 200 ng of total RNA was used as input for the Illumina TotalPrep-96 RNA amplification kit (cat: 4393543, based on the method published by the Eberwine lab (19) to generate and bead-purify cRNA amplicons. The amplified cRNA was normalized to 150 ng in 5 µL of nuclease-free water (or via speed-vac if the sample was too dilute) and hybridized to Illumina Human HT-12 bead-chips (v. 4.0, cat: BD-103-0604) in accordance with the Illumina "Whole-Genome Gene Expression Direct Hybridization Assay Guide" (revision A). Briefly, cRNA was added to the prepared arrays and incubated overnight (approximately 14-16 hours) in an Illumina Hybridization Chamber / oven at 58°C with constant gentle rocking. Following hybridization, the chips were washed, blocked, and dried via centrifugation before being imaged on an Illumina iScan system. An .idat file was produced for each of the scanned arrays, which was then exported for downstream analyses.

### Methylome Profiling

Baseline or screen samples (when baseline was not available) were randomized to ensure that the distributions of participant demographics were evenly represented across each 96-well batch. Each sample was quantified using the Quant-iT PicoGreen dsDNA Assay Kit (ThermoFisher, CAT: P7589) with a plate-reader using a standardized curve. The samples were normalized to 500 ng in 45 µL of nuclease-free water before being bisulfite treated using a Zymo EZ DNA Methylation kit (catalog number: D5001; Zymo Research, Irvine, CA) using PCR conditions for Illumina's Infinium Methylation assay (95°C for 30 seconds, 50°C for 60 minutes×16 cycles). Methylation data was generated using the Illumina MethylationEPIC Beadchip (catalog number: WG-317-1002; Illumina, San Diego, CA) following manufacturers protocols. Briefly, a total of 4 μL of bisulfite converted DNA was hybridized to Illumina BeadChips. Samples were denatured and amplified overnight for 20 to 24 hours. Fragmentation, precipitation, and resuspension of the samples followed overnight incubation, before hybridization to EPIC BeadChips for 16 to 24 hours. BeadChips were then washed to remove any unhybridized DNA and labeled with nucleotides to extend the primers to the DNA sample. Following the Infinium HD Methylation protocol, the BeadChips were imaged using the Illumina iScan system (Illumina). The completed run files were exported in .idat format and used for downstream analyses.

### Targeted OXTR Methylation

All viable DNA samples available were included in targeted pyrosequencing. Samples were quantified on a multi-channel spectrophotometer prior to being normalized to a concentration of 12.5 ng/µL (500 ng total) in a 96-well PCR plate. The bisulfite reaction was performed using the EpiTect Fast 96-well format conversion kits (Qiagen, CAT:59720) using the "low concentration" version of the standard protocol for DNA samples (published 06/2012). The following extended-incubation version of the conversion program was used on a 96-well thermal cycler to ensure complete conversion. Following the bisulfite conversion, samples were purified and eluted in 70 µL of Qiagen elution buffer (CAT: 19086). The bisulfite-converted DNA was then stored at 4°C prior to downstream PCR amplification, or at -20°C if the samples would not be used in the next 24 hours.

Biotinylated PCR primers and a nested sequencing primer were designed to target three CpG sites within the MT2 region of OXTR. These sites were selected based on the findings of Gregory *et al.* (16) that identified association between hypermethylation of several CpGs in the promoter region of *OXTR* with ASD in both peripheral blood and post-mortem temporal cortex samples.

DNA sequence of target region plus an additional 200 bp of flanking DNA was used to generate a list of candidate PCR and sequencing primers using the Pyromark Assay Design software (v 2.0.1.15, Qiagen, MD). PCR primer pairs were selected based on compatible melting temperatures and estimates of secondary structure formation, ambiguous local mapping, or self-annealing (**Supplementary Table 1**)**.** PCR reactions consisted of 2 µL of bisulfite-converted DNA and 23 µL of Pyromark PCR Master Mix (Qiagen, CAT:978703). Each sample was run in triplicate on a 96-well PCR plate

Prior to sequencing, 5 µL of each post-PCR sample was added to 75 µL of bead buffer containing 1.5 µL of Streptavidin Sepharose High Performance beads (GE Healthcare, CAT: 17-5113-01). The samples were placed on a shaker set to 1,400 rpm for at least 10 minutes, extracted from the solution using a 96-prong vacuum station, and cycled through reservoirs of 70% ethanol, denaturation solution (Qiagen, CAT: 979007), and 1x wash buffer (Qiagen, CAT: 979008). The vacuum was then turned off to release the beads into a 96-well clear-bottom plate containing the annealing solution and the sequencing primers. The sequencing plate was heated on a pre-warmed block set to 80°C for two minutes and allowed to return to room temperature before proceeding. The bead-bound amplicons were sequenced on a Q96 pyrosequencer using PyroMark Gold Q96 CDT Reagents (Qiagen, CAT: 972824) loaded into PyroMark Q96 HS Capillary Tips (CDT) (Qiagen, CAT: 979104) (**Supplementary Table 5**)

### Quality Control Procedures

#### Genome-Wide Genotyping

Initial quality control procedures were executed in Illumina’s GenomeStudio software. First, All Y chromosome markers, all X chromosome markers with more than 2 male heterozygous calls, and all SNPs with a call frequency less than 98% were set to zero. SNPs were also set to zero if they had an AB or AA R Mean (normalized intensity of heterozygous clusters) value of < 0.20, or if AB T Mean (normalized theta values of heterozygous clusters) fell within the following ranges: [0>= AB T Mean >= 0.2, 0.8 >= AB T Mean >= 1.0]. SNPs were manually reviewed if they had an AB T Mean value within the following ranges: [0.75 <= AB T Mean < 0.80, 0.2 < AB T Mean <= 0.25]. Subsequently, SNPs were zeroed if they had a cluster separation metric of < -0.30, and SNPs with cluster separation metrics of > 0.30 and < 0.35 were manually reviewed. Manual reviews were also performed for SNPs demonstrating abnormally low or abnormally high heterozygosity [0.2 <= Het Excess <= 1.0, -1 <= Het Excess <= -.03]. Within these samples, SNPs with AA or BB R dev values >= 0.05 or AB T dev values >= 0.05 were reviewed and SNPs with AA or BB T dev values >= 0.05 were zeroed. Finally, all SNPs with > 3 replication errors or < 0.98 call rate were zeroed.

The data were then exported and converted into a format compatible with PLINK (v1.9, (20)) which was used for the majority of all subsequent quality control steps. Although the genotypes were originally exported from GenomeStudio using the hg38 build, to have consistency across all datasets, probe coordinates were reverted to the hg19 build and annotations from the InfiniumOmni2-5-8v1-3_A1 manifest and its companion files were used to flag and remove probes with overlapping positions and/or being categorized as "unmapped" or "in-del" probes (n = 11,405). If available, each probe ID was converted into its respective rsID using the InfiniumOmni2-5-8v1-3_A1_b144_rsids manifest, and two additional probes were dropped from the Y chromosome, which was not included in downstream genotypic analyses. It should be noted that X chromosome probes, as well as those annotated to XY (pseudo-autosomal) and M (mitochondria) were retained in the dataset for quality control procedures.

Samples were flagged and removed if their predicted sex conflicted with their study-annotated sex (n = 3). Two of these belonged to participants with known sex chromosome aneuploidies, while the third reflected a likely sample swap issue specific to that sample. Probes were removed if they were flagged for Mendelian errors (identified using one two-parent, two-child CEPH pedigree genotyped alongside study samples for quality control purposes) or if there was any discordance between samples from the same individual in our technical replicates (n = 1,022). Additionally, heterozygous haploid genotype calls were assumed to be genotyping errors and set to missing. In instances where genotypes were available for more than one array per participant, the array with the higher rate of missing genotype data was preferentially excluded (n = 10 excluded).

In order to determine whether our study samples deviated significantly between their self-report race and their genotypically determined ancestry, which could have potentially reflected a sample swap, we downloaded publicly available HapMap Phase 3 genotypes. In parallel with our SOARS-B samples, HapMap 3 autosomal genotypes were filtered to SNPs only and a MAF of 0.05, and samples were restricted to a missing rate < 0.1. Non-ambiguous SNPs present in both datasets were retained (i.e. A/T and C/G SNPs were excluded), and after excluding non-founders, two rounds of LD-pruning were applied within the HapMap3 dataset only (using parameters --indep 50 5 2 and --indep-pairwise 500 50 0.2, respectively). The SOARS-B dataset was then restricted to the remaining independent SNPs from HapMap 3, strand consistency was enforced, and the datasets were merged. Principal Components Analysis (PCA) using EIGENSOFT's smartpca function (21, 22) was then performed, where 20 PCs, with no outlier removal, were calculated only within HapMap 3 samples and projected onto SOARS-B samples. From the resulting PCs, we confirmed that all samples roughly matched their self-reported race, when available.

Probes were further filtered by removing any with a calculated missingness >= 0.01 in the study population (n = 36,489) and any probes that were monomorphic (n = 241,184). Non-SOARS-B samples (i.e. six DNA samples purchased through Coriell, including the four-person CEPH pedigree and a pair of unrelated African-American samples) were then removed from the dataset. To calculate homozygosity (where significant deviations can indicate larger, more substantial genomic aberrations that wouldn't be generalizable in a population setting) and genetic relatedness amongst samples, common variants (MAF >= 0.01) were pruned for linkage disequilibrium and restricted to autosomes-only (using LD-pruning parameters identical to those used with the HapMap 3 dataset). These quality checks identified one pair of individuals related at a pi-hat > 0.185, where the individual with fewer available study samples was excluded from further analysis, and one individual who was an outlier (> 6 standard deviations away from the mean of all samples) for homozygosity and was also excluded from further analysis. After all participant filtering steps were completed, a final variant-level MAF and missingness filter was applied that excluded an additional 5,640 probes. In totality, this quality control procedure resulted in a final dataset consisting of 281 individuals and 2,049,113 probes.

Using this quality-controlled dataset, PCA using EIGENSOFT was run again on common (MAF 0.01), LD-pruned autosomal variants and used to calculate principal components that were used in downstream association testing to control for genetic ancestry.

#### Targeted Genotyping

Initial quality control of the n=12 TAQMAN SNPs was applied using ViiA 7 software (ThermoFisher, MA) to identify and remove ambiguous genotype cluster assignments. One sample failed initial and repeat genotyping with the rs4813627 TAQMAN assay and was not included in the analysis. As a rough approximation of sample identity confirmation, participants’ X-chromosome genotypes were compared to those generated on the Omni 2.5 to confirm genotype agreement. Subsequent sample-level quality control was applied as described for the genome-wide analyses and resulted in a dataset containing n = 281 samples across n = 12 SNPs.

#### CNVs

CNVs were called on all genotyped samples using the PennCNV software (23-25). For generating CNV calls, we utilized the Hidden Markov Model file hhall.hmm supplied by PennCNV, a Population B Allele Frequency (“pfb”) file which set the PFB for every probe to 2, i.e. treating each probe as intensity-only, and a GCModel file, which helps control for GC content during CNV calling, using the same method as described in the supplemental methods of Szatkiewicz et al (26). The decision to treat probes as intensity-only stemmed from our relatively small sample size, ancestrally non-homogenous study population, and lack of ability to generate pfb files for participants of non-European ancestry. Quality control procedures were heavily influenced by those applied in Marshall et al. (27) and were as follows: autosomal and X chromosome CNVs were called using the input files as described above. For both autosomal and X chromosome CNVs, low quality CNVs (<10 SNPs, confidence score <10 & <10kb in length) were removed and then samples were excluded if they were > the median + 3 standard deviations for the following metrics: LRRSD, BAFSD, absolute GCWF value, number of autosomal CNV calls, or total autosomal CNV length, with both number of CNVs and total length being calculated after removing low-confidence CNVs. CNVs in the remaining samples were then annealed (based on probes) using the clean_cnv.pl function and –fraction 0.2 parameter. Using the scan_region.pl function, CNVs were then further filtered by excluding those that overlapped >= 50% with a genomic gap region (--minqueryfrac 0.5 parameter) or those that reciprocally overlapped >= 50% with a segmental duplication region (--minoverlap 0.5 parameter). Both genomic gap and segmental duplication region coordinates in hg19 were obtained via the UCSC table browser. For pairs of samples that belonged to the same participant, the sample with higher post-QC median autosomal CNV confidence score was retained & the other was excluded from further analysis. Non-study samples and samples flagged for exclusion using the Omni 2.5 data were then excluded. X-chromosome CNVs were also not considered further for analysis due to the small number of post-QC calls (n=22). Autosomal CNVs were then further filtered to be a minimum of 20kb in length.

To determine the quality of our CNV calling for the CNVs considered for analysis, for individuals that were genotyped twice and had post-QC CNV calls available, concordance of autosomal CNV calls across the samples was calculated at varying levels of minimum reciprocal overlap for defining concordance. At 100%, 80%, and 50% reciprocal overlap thresholds, the median concordance rates were 83%, 87% & 87% respectively. Of a pair of samples where there was particularly low concordance (25% for all thresholds), the sample with the fewer number of CNV calls had already been selected for retention in the dataset.

After all quality control procedures and filtering CNVs to >= 20kb length, 259 participants had a total of 1,198 autosomal CNVs. CNVs ranged from 20-952kb in length, with a median length of 36kb, and 46% were duplications.

#### Transcriptome

Each .idat file obtained from the iScan corresponded to one biological sample from the SOARS-B study. We excluded any sample with a signal-to-noise ratio <= 2 (n = 48). Detection p-values were calculated for each probe across all samples in each batch and samples were removed if: 1 - their proportion of expressed probes was less than three standard deviations from the batch mean; and 2 - their median expression values were less than or greater than three standard deviations from the batch mean (n = 32). All four batches were then combined, background corrected, and quantile normalized using the neqc function within limma. Sex discrepancies were assessed using the mean expression levels of X- and Y-associated probes, which led to the removal of one sample with an annotated sex that conflicted with its predicted sex. Identity conflicts across longitudinal time points were assessed using the ILMN-2399463 probe, which overlaps with the SNP rs8676, leading to a genotype-specific change in expression that can be used to identify mislabeling or sample swaps (28). This approach flagged one additional sample that was removed from the analysis. Eleven samples were dropped due to flags in the population genetics analyses, which are detailed in the genotype array QC pipeline. Finally, all control samples and technical replicates were dropped from the analysis, resulting in a final dataset of 809 samples.

At the probe level, all probes that were flagged as "Bad" or "No match" for the probe's target mapping quality from the Bioconductor package "illuminaHumanv4.db" (29) were removed from the analysis (n = 12,847). Additionally, probes were dropped if they were not detected in any sample using a detection p-value < 0.05 (>= 0.05 was considered non-detected; n = 2,843) or if they were flagged due to issues identified after their initial design (n = 3,261). Probes annotated as overlapping SNPs had their minor allele frequencies (MAF) queried using Ensembl and were excluded if any overlapping SNP had a MAF > 1%, as well as probes whose sequences were annotated as mapping to >1 location in the genome. Collectively, these filtering parameters led to the inclusion of 24,002 probes in the cleaned dataset.

#### Methylome

Raw .idat files from the EPIC methylation arrays were downloaded from the Illumina iScan and imported into R as a red-green channel dataset using the "minfi" package (v 1.36, (30)). Cell proportion estimates were generated for each sample using the "estimateCellCounts" function in minfi with the blood cell reference dataset and appended to the phenotype annotations for each sample. This function generates proportional estimates of CD8 T cells, CD4 T cells, natural killer cells, B cells, monocytes, and granulocytes based on the method developed by Houseman *et al.* (31). Background correction was performed using noob (32) and predicted sex was calculated. Five arrays (belonging to n = 3 unique biological samples/participants, two of which had two technical replicates present in the dataset) were identified as having discrepant methylation-predicted vs. self-reported sex. These discrepancies were identical to those observed in the genotype data; three of the discordant arrays (representing two samples) reflected the n = 2 participants known to have sex chromosome conditions, and two arrays (one sample) reflected the likely sample-swap issue previously observed. In parallel, the raw .idat files were read in using the "ewastools" package (33) to check whether samples passed Illumina's quality control thresholds (n = 7 were flagged for failing Bisulfite II conversion, but not excluded from analyses) and to confirm genotype agreement (called from methylation signals) between technical replicates. Probes' values were set to missing if the detection p-value was >= 0.01. Probes were removed from the dataset if they had missing data for >= 1% of the study population (n = 4,515), and samples were removed if they were missing >= 5% of the post-QC probes (n = 0). Nine arrays representing technical replicates were also excluded from the dataset, leaving n = 272 unique samples. Finally, probes were excluded if they were within 2 base pairs of a SNP with a MAF >= 0.05, as determined by the rmSNPandCH function within R package DMRcate (v 2.4.1, (34)) (n = 58,503 probes). This QC pipeline led to a final quality-controlled dataset consisting of 802,841 probes and 272 individuals, 173 of whom had complete data for all subsequent model variables.

#### Targeted OXTR Methylation

Three PCR-replicates were generated from each bisulfite-converted DNA sample. This approach was used to address inherent variability in PCR amplification efficiency of methylated and bisulfite-converted DNA, which is the primary source of variation in methylation estimates from pyrosequencing data. Pyrograms of each reaction were saved and the raw text-outputs of the Q96 analysis software were imported into R. The following QC pipeline was used to filter out results with variation that exceeded the 5% limit.

When possible, the original DNA aliquot was used to rerun the samples that failed QC filtering. In some cases, samples had been depleted, and reallocations for the same timepoint were necessary, but the protocol for each of the rerun samples was identical to those previously described. One sample was excluded for exhibiting extreme hypomethylation across all 3 CpGs.

### Data Analysis and Supplementary Results

**Pathway Analysis**

Probe level results from the gene expression and DNA-methylation arrays were used as inputs for the fGSEA analysis. Analyses were restricted to a single probe for each gene in each dataset. If multiple probes were available for a gene, the probe with the lowest p-value was selected. Enrichment scores were calculated using log2 fold change (log2FC) values across five gene sets (MSigDB: Hallmark, Gene Ontology (GO): Biological Process, GO: Cellular Component, GO: Molecular Function, and MSigDB: KEGG). Results were obtained by running fGSEA with the adaptive multilevel splitting Monte Carlo method (fgseaMultilevel) and an eps (minimum p-value) of 0. The collapsePathways function was used to minimize redundancy between pathways in each gene set and an adjusted p-value of 0.1 was used to define significant enrichment.

#### Targeted Genotypes

Eleven of the twelve targeted SNPs were tested for association with oxytocin. The X-chromosome SNP was not included for association testing as its purpose was solely to confirm genotype agreement across our targeted and genome-wide assays.

#### CNVs

Previous studies have shown that rare CNVs can play a role in the etiology of ASD and thus we incorporate an assessment of CNV burden testing in our analysis (35). To perform rare CNV burden testing, we took two approaches in parallel, CONCUR (36), a locus-free kernel-based algorithm for CNV burden testing, and basic linear modeling in R. For both approaches, CNVs were subset into 3 datasets based on copy number type: deletions & duplications together, deletions-only and duplications-only. Including only individuals with all analysis covariates, CNVs were then frequency filtered using a previous version of PLINK with the parameters --cnv-freq-exclude-above 7 --cnv-overlap 0.5 (roughly a 5% frequency filter based on the n = 153 individuals with at least one CNV and all analysis variables, with a 50% overlap definition). In both analyses, included covariates were functional strata, sex, 3 ancestry PCs, age at sample, and oxytocin measurement batch. For the linear model association testing, CNV burden was modeled as total number of CNVs and as total length of CNVs in kb.

#### Multi-Omic

In order to simultaneously leverage our three ‘-omic’ datasets, we used an integrative approach called Multi-Factor Omics Analysis + (“MOFA+”), which was developed for integrating and analyzing multiple biologically-relevant datasets taken on a common core of study participants. Because of author recommendations that MOFA+ “views” (i.e. individual datasets) are approximately similarly sized in terms of the number of features, we decided on a target set of roughly 8,500 genes as input from each dataset, determined by the number of features available in our gene expression dataset, which only retained 8,309 genes when we restricted to probes expressed in >50% of the samples. For gene expression and methylation array data, the most variably expressed/methylation (using M values) probe/CpG was kept per gene and, to reduce the impact of confounders, analyzed in a linear model using limma, adjusting for sex, study site, ancestry principal components and gene expression batch or sex, study site, ancestry principal components, cellular composition variables, sample plate and array row, respectively. The residuals from these linear models were then used as input for MOFA+ factor creation. SNPs of samples which passed genotype QC were filtered to a minimum minor allele frequency of 1%, LD pruned (using the same parameters for PC construction) and intersected with genic regions. Genic regions were obtained via biomaRt’s Ensembl GRCh37.p13 reference, and only genes with a HGNC ID and annotated to autosomes, sex chromosomes or the mitochondria were retained. Genes with a gene biotype classified as ‘psuedogene’ were excluded, and, as with other genic regions utilized in this paper, 5kb was added to either side of the coordinates. One SNP per gene was randomly selected and its genotype calls retained as input for MOFA+. Using these three datasets as input into MOFA+, we applied within-dataset scaling and utilized the slow convergence mode to construct 20 factors. All samples with available post-QC data in at least one of the views were included in factor creation.

While we had anticipated that the resulting factors might reflect roughly equal variances from each of the input views, each of our factors was dominated by one view, with very little of the other two view’s variance being explained by the factor. Furthermore, relatively very little of the genotype and methylation views’ variance were captured by the 20 resulting factors (<20% variance each), while >60% of the gene expression view’s variance was explained.

The resulting factors were then tested as independent variables of interest for association with pre-treatment oxytocin, in univariate linear models and linear models adjusting for oxytocin measurement batch. For factors that had a nominal association with plasma oxytocin (p-value<0.05) in models adjusting for oxytocin measurement batch, we utilized MOFA+’s run_enrichment function to get the top 3 KEGG pathways for positive and negative loadings of the most dominant view separately

10.	Dan T (2020): KEGGREST: Client-side REST access to the Kyoto Encyclopedia of Genes and Genomes (KEGG). 1.30.1.

29.	Dunning M LAEM Illumina HumanHT12v4 annotation data (chip illuminaHumanv4). *Bioconductor*.
