## Supplementary Figures for "Genetic and epigenetic signatures associated with plasma oxytocin levels in children and adolescents with autism spectrum disorder"

Supplementary Figure 1: Pre-treatment nat. log-transformed plasma OXT by study site.

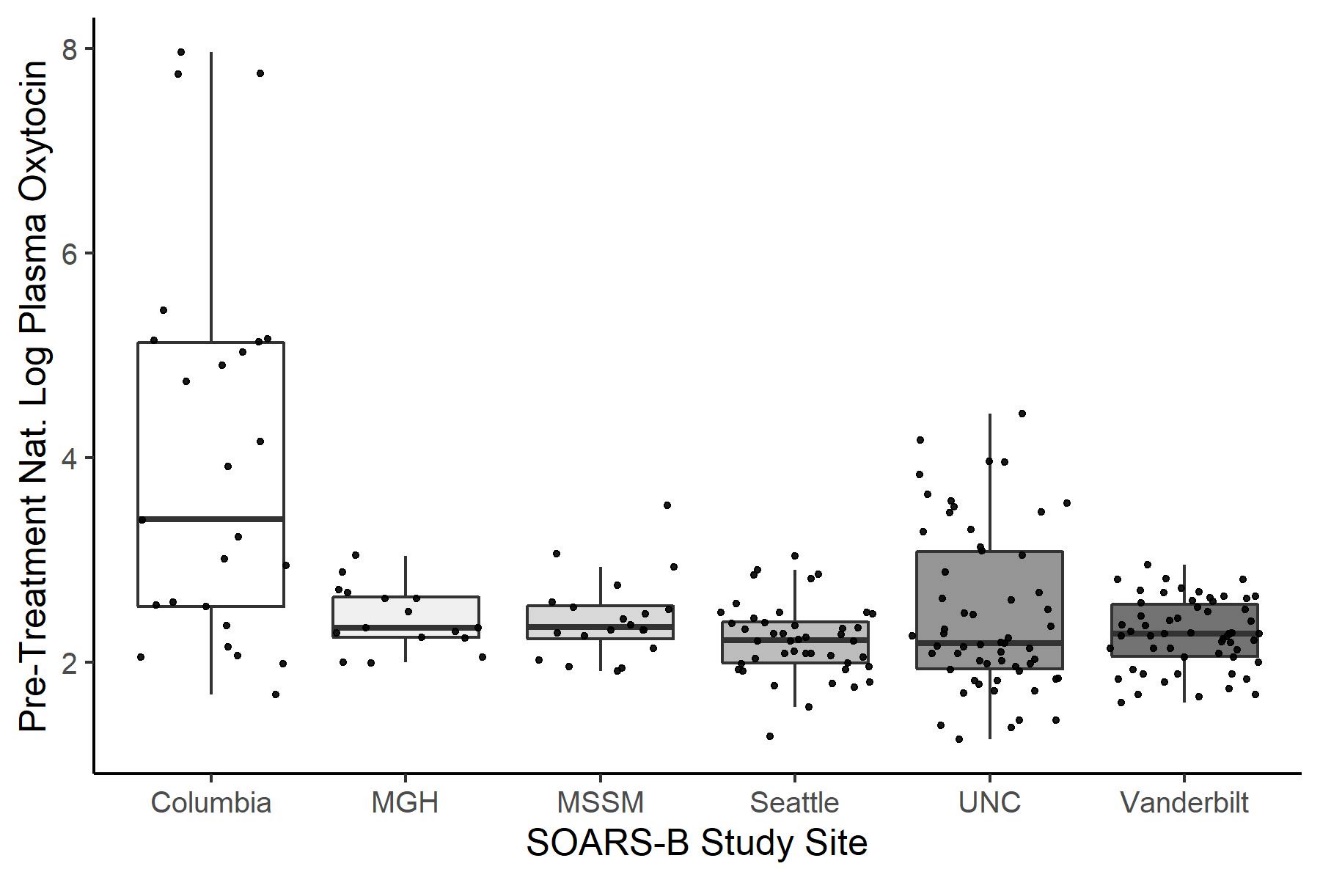

Supplementary Figure 2: Boxplot for top GWAS hit, rs6500746, within RBFOX1 with plasma oxytocin

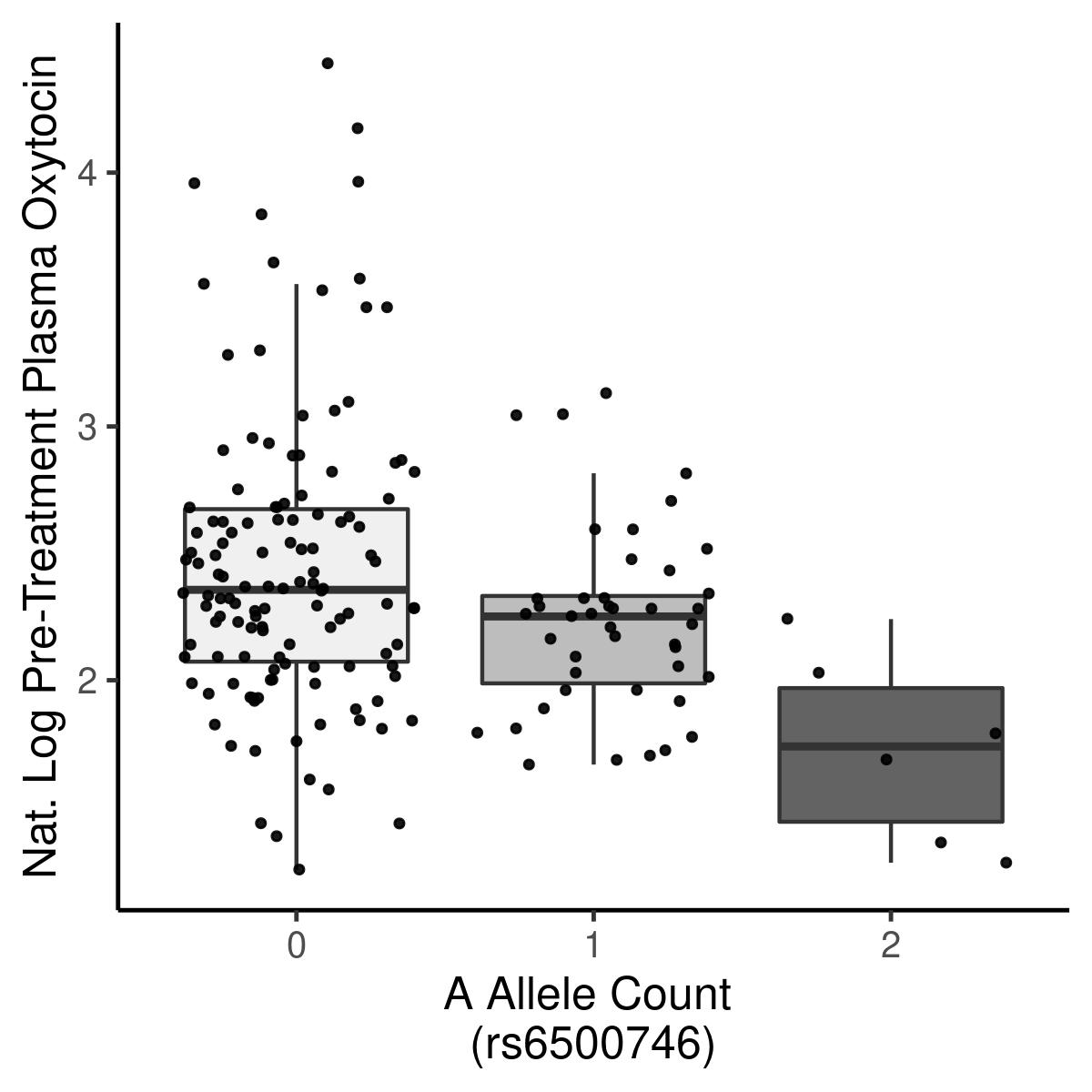

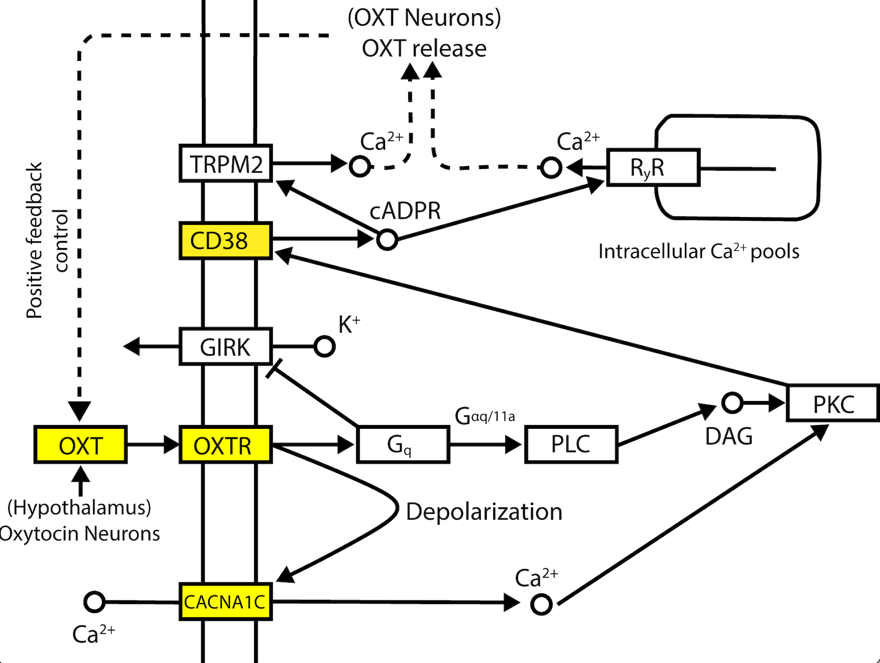
Supplementary Figure 3: KEGG annotated oxytocin signaling pathway for Taqman assays
